# Volume and Distribution of White Matter Hyperintensities in Rheumatoid Arthritis and Ulcerative Colitis Patients

**DOI:** 10.1101/2024.05.30.24308189

**Authors:** Jennifer G. Cox, James H. Cole, Matthew J. Kempton, Steven C. R. Williams, Marius de Groot

## Abstract

Brain white matter disruptions have been implicated in contributing to fatigue, brain fog and other central symptoms commonly reported in inflammatory diseases. In this study, we included 252 RA patients with 756 age and sex matched controls and 240 UC patients with 720 age and sex matched controls using the UK Biobank imaging dataset. We looked for differences in total volume of white matter hyperintensities (WMH) between patients compared to controls. Then, using voxelwise analysis, we explored the spatial distribution of these white matter hyperintensities and differences in these between patients and controls and between disease groups.

A significantly higher volume of WMH was observed in both the RA (p = 2.0 x 10^-8^, β = - 0.36, 95% CI = -0.48, -0.23) and UC (p = 0.003, β = -0.19 95% CI = -0.32, -0.07) patients compared to their respective control groups. Voxelwise analysis revealed only a small cluster of RA associated WMH compared to controls.

These results indicate an increased risk of white matter hyperintensities in patients with RA and UC. These findings help quantify the effect of inflammation from autoimmune diseases on cerebrovascular health and white matter integrity.

## Introduction

Fatigue is a prominent feature of autoimmune diseases and is a large area of focus in rheumatoid arthritis (RA) and ulcerative colitis (UC) research [1, 2]. There is an extensive body of previously published work exploring neuroimaging in these chronic inflammatory diseases looking to identify surrogate biomarkers of disease linked to fatigue [3]. Brain white matter disruptions in the form of changes to white matter microstructure and white matter lesions (WML) have been implicated in contributing to fatigue, brain fog and other central symptoms commonly reported in inflammatory diseases [4].

The presence of WML is strongly associated with atherosclerosis [5]. Vice versa, both RA and UC are associated with higher levels of atherosclerotic cardiovascular diseases (ASCVD). Therefore, even when controlling for traditional cardiovascular risk factors, higher levels of accelerated atherosclerosis are seen in both these diseases and in other chronic inflammatory diseases. One of the primary theories on this increased risk is due to shared pathology between these diseases, including changes in the innate and adaptive immune systems resulting in systemic inflammation and endothelial dysfunction [5-8].

WMHs present as high signal intensity areas on T2-weighted or fluid-attenuated inversion recovery (FLAIR) MRI sequences. They are indicative of changes in white matter fibers and tracts [9]. There are several studies that look specifically at brain white matter in both RA and UC [10, 11]. However, the literature around white matter hyperintensities has been conflicting. One of the earliest papers to look at WMHs in RA, Bekkelund et al., did not show a significant difference in WML load between RA patients and controls [12]. A more recent study was published by Vassilaki et al. using the Mayo clinic study of aging cohort that did report a significant difference in WMH volume in RA patients [13]. There are key analytical design differences between these two papers. The Bekkelund et al. paper analysed lesion count and the area of the largest identified lesion, whereas Vassilaki et al. measured total WML volume as a function of total intracranial volume (TIV). This more recent work provides a view of the total average lesion load in RA patients compared to controls.

These mixed reports are further complicated by our lack of understanding of the role of these lesions in disease processes. There is a significant correlation between the presence of these lesions and age and can be seen in otherwise healthy populations [14, 15]. Conversely, there is a large body of evidence which suggests that these lesions can be linked to various different diseases and inflammatory processes and are associated with cognitive decline and increased risk of dementia [16-18].

The aims of this study were to determine if there were differences in the presentation of white matter hyperintensities between patients with RA compared to matched controls and patients with UC compared to matched controls in the UK Biobank population. As evidenced by the current published data there are different methods for analysing WMH load and distribution. There are various visual scoring methods (Fazekas, Scheltens scale and the Age-Related White Matter Changes scales) available [19-21]. It is also not uncommon to identify a primary lesion for comparison. However, in this study we have the benefit of a very standardised data set and so we chose to look at total lesion volume using binary lesion masks acquired using a fully automated, supervised-learning method of WMH identification. The primary analysis focused on the total volume of white matter hyperintensity (WMH) differences between groups with a follow-up analysis looking at differences in the distribution of these WMHs. Additionally, we looked to see if there was a difference in the distribution of these WMHs between these two diseases directly.

## Methods

### Study population

This is a case control study utilising the UK Biobank dataset. The UK Biobank is a large, prospective observational study of 500,000 participants providing extensive biological information [22]. The imaging substudy is planned to scan 100,000 of those participants with a standardised scanning protocol including MRI of the brain.

At the time of this investigation, brain MRI were available from 40,681 participants. For the purposes of the present study, we selected 1,968 individuals including patients with RA, UC and healthy controls. Due to the difference in age and sex distribution between RA and UC patient populations, separate control groups were matched to each patient population in a 1:3 patient:control ratio. Using the matchit algorithm in R, an exact matching strategy was employed for sex and a nearest neighbour matching strategy was utilised for age matching and selection of the control groups. A matching ratio of 1:3 was determined to be optimal as it allowed for the highest matching ratio and so statistical power, while utilising the matching strategy outlined above.

For the initial analysis looking at total WMH volume, data from 252 individuals with a primary or secondary diagnosis of RA identified using International Clarification of Disease (ICD)-10 codes M05 or M06 were included in the RA patient group (mean age ± SD in RA = 65.52 ± 7.03, 71% female) with 756 age and sex matched controls (mean age ± SD in RA control group = 65.52 ± 7.02, 71% female). Data from 240 individuals with a primary or secondary diagnosis of UC identified using ICD-10 code K51 were included in the UC patient group (mean age ± SD in UC = 63.98 ± 7.06, 51% female) with 720 age and sex matched controls (mean age ± SD in UC control group = 63.98 ± 7.05, 51% female).

For the follow-up analysis looking at distribution of WMH data from 207 individuals with a primary or secondary diagnosis of RA identified using International Clarification of Disease (ICD)-10 codes M05 or M06 were included in the RA patient group (mean age ± SD in RA = 65.82 ± 7.13, 70% female) with 207 age, sex and total volume of white matter hyperintensity IDP matched controls (mean age ± SD in RA control group = 67.47 ± 6.67, 69% female). Data from 211 individuals with a primary or secondary diagnosis of UC identified using ICD-10 code K51 were included in the UC patient group (mean age ± SD in UC = 64.31 ± 7.00, 51% female) with 211 age, sex and WMH IDP matched controls (mean age ± SD in UC control group = 64.50 ± 7.04, 47% female). These individuals are a subset of the subjects used for the total WMH volume analysis based on the availability of the lesion maps. Further detail on the lesion map availability can be found in the following section on data acquisition and processing.

### Data Acquisition and Processing

Multi-modal MR images were acquired on Siemens Skyra 3T scanners running VD13A with a standard Siemens 32-channel RF receive head coil. The T1-weighted MRI used an MPRAGE sequence with 1-mm isotropic resolution, 256×256×208 matrix, T1/TR = 800/2,000 ms, R=2. The T2-weighted MRI was a structural FLAIR acquisition with 256x256x192 matrix, TI = 1,800 ms, TR= 5,000 ms, TE=395 ms, R=2 and elliptical k-space coverage. The T2-FLAIR image was linearly aligned to the T1-weighted image using FSL FLIRT [23, 24]. Lesion segmentation was automatically carried out using the BIANCA (Brain Intensity AbNormality Classification Algorithm) tool [25]. BIANCA is a fully automated method for white matter hyperintensity detection and classification based on the k-nearest neighbour algorithm. BIANCA classifies image voxels based on their intensity and spatial features. The result is a binary mask output image which represents the probability per voxel of being WMH. From this mask an imaging derived phenotype (IDP) is generated for each participant for total volume of WMHs.

The output from BIANCA is a probability map of voxels to be classified as lesions. The interim step between this lesion identification and the total volume of WMH IDP is a binary lesion mask. To create a summary lesion map for the UC and RA patient populations these individual masks were first transformed from their native T1 space to standard MNI152 space. The individual warp coefficients were provided as part of the UK Biobank pre-processing pipeline. This file was then applied to the individual lesion masks. A summary image of all subjects within each patient population was created by concatenating all images as a time series. This 4D output image was then mean filtered to create a single average image for each patient and control population.

As a next step to look at the distribution of these white matter hyperintensities by population, summary lesion maps for RA and UC along with their respective control groups were created from the binary lesion masks. The lesion segmentation analysis was not added to the analytical pipeline until January 2017. Scans acquired before this date were processed through the BIANCA pipeline and the summary IDP from that output has been provided. However, any subjects that were scanned and released prior to this date do not have a lesion mask available to download and patients scanned before this date were therefore excluded from the voxelwise analysis.

Given this limitation to the lesion map availability the lesion distribution analysis was performed using a 1:1 matching strategy between the patient and control groups. The same control group that was originally sampled was used with an additional matching performed based on total volume of WMH IDP.

Full details on the UK Biobank neuroimaging data are provided here: https://biobank.ctsu.ox.ac.uk/crystal/crystal/docs/brain_mri.pdf

### Statistical Analysis

All statistical analyses were carried out using R version 4.1.1 and FSL version 6.0.5.2

### Total Volume of WMH

The initial analysis for this study compared total volume of white matter hyperintensities in each patient group to their respective matched control groups. Given the non-normal distribution of these volumes all analyses were done using the log transform of the ratio of total volume of white matter hyperintensities to total brain volume. Model one consisted of a linear model regression with sex, age and intracranial volume (ICV) as covariates.

Hypertension is a known risk factor for the presence and burden of white matter hyperintensities in subcortical and periventricular regions of the brain [26, 27]. Given this association and the increased prevalence of hypertension in both the UC and RA patient populations, as reported in the demographics table below, a second model was run analysing total volume ratio of WMHs with hypertension as an additional covariate to sex, age and ICV. This is also in keeping with previously run analyses.

To calculate effect sizes, we used the regression (β) coefficient. All two-sided p-values < 0.05 were considered statistically significant.

### Voxelwise Analysis

As a next step, voxelwise statistical analysis was performed using Randomise, FSL’s tool for nonparametric permutation-based testing [28]. The aim of this analysis was to explore differences in the spatial distribution of these white matter hyperintensities. Three separate two sample paired t-test analyses were performed: RA compared to controls, UC compared to controls and RA subjects compared to UC subjects.

The analyses were run with 10,000 permutations. Given the difference in variable distributions between the groups all data was demeaned. The Threshold-Free Cluster Enhancement (TFCE) method was used and provided a test statistic to identify significant clusters. Utilising the atlasquery function in FSL the max and min coordinates for any clusters were run against two separate atlases, the Harvard-Oxford Subcortical Structural Atlas and the JHU ICBM-DTI-81 White-Matter Labels atlas [29-31].

## Results

### Demographics

Detailed demographic information can be found in Table 1. The control groups were matched directly on age and sex. Both control groups were generally comparable to their matched patient population except for rates of hypertension and hypercholesterolemia. For the purposes of this analysis hypertension and hypercholesterolemia were defined based on the ICD-10 codes for these diseases. As this discrepancy was anticipated, we examined the role of hypertension in these diseases using a second model with hypertension as a covariate.

**Table 1:** Participant demographics

|  | RA<br>n=252 | RA<br>Controls<br>n=756 | p value<br>(RA vs<br>RA<br>Control<br>) | UC<br>n=240 | UC<br>Controls<br>n=720 | p value<br>(UC vs<br>UC<br>Control<br>) |
| --- | --- | --- | --- | --- | --- | --- |
| Sex | 180 F /<br>72 M | 540 F /<br>216 M | 0.99 | 122F<br>/118 M | 366 F<br>/354 M | 0.99 |
| Age (y ± SD) | 65.52 ±<br>7.03 | 65.52 ±<br>7.02 | 0.99 | 63.98 ±<br>7.06 | 63.98 ±<br>7.05 | 0.99 |
| ICV (mL) | 1155 ±<br>113 | 1161 ±<br>112 | 0.77 | 1189 ±<br>117 | 1196 ±<br>116 | 0.89 |
| Hypertension % (n) | 33 (84) | 16 (118) | <0.001 | 29 (70) | 17 (121) | <0.001 |
| Education - %<br>subjects with a<br>college/university<br>degree or professional<br>qualification (n) | 43 (109) | 49 (372) | 0.06 | 46 (111) | 51 (370) | 0.09 |
| Diabetes % (n) | 7 (17) | 4 (30) | 0.13 | 9 (21) | 5 (35) | 0.04 |
| Smoking Status |  |  |  |  |  |  |
| Current % (n) | 9 (22) | 5 (39) | 0.03 | 2 (5) | 7 (50) | 0.01 |
| Previous % (n) | 40 (102) | 35 (264) |  | 43 (102) | 35 (249) |  |
| Never % (n) | 51 (128) | 60 (453) |  | 55 (133) | 58 (421) |  |
| Ethnicity % white (n) | 96 (241) | 98 (741) | 0.03 | 95 (229) | 98 (704) | 0.03 |
| Hypercholesterolemia<br>% (n) | 18 (46) | 9 (65) | <0.001 | 11 (26) | 8 (59) | 0.11 |

Variables listed as % (number of participants). RA stands for rheumatoid arthritis, UC ulcerative colitis, SD standard deviation, ICV intracranial volume.

### Total white matter hyperintensity volume in RA and UC

A significantly higher total volume of WMHswas observed in both the RA (p = 2.0 x 10^-8^, β = -0.36, 95% CI = -0.48, -0.23) and UC (p = 0.003, β = -0.19 95% CI = -0.32, -0.07) patients compared to their respective control groups. Given the non-normality of the data the linear model regression was run using the log transform of the ratio of total WMH volume to total brain volume. This allows for a normalisation on a group level and on an individual level given expected differences in individual total brain volume. The results for total volume of WMHs in RA can be found in Figure 2. The results for total volume of WMHs in UC can be found in Figure 3.

**Figure 2.**
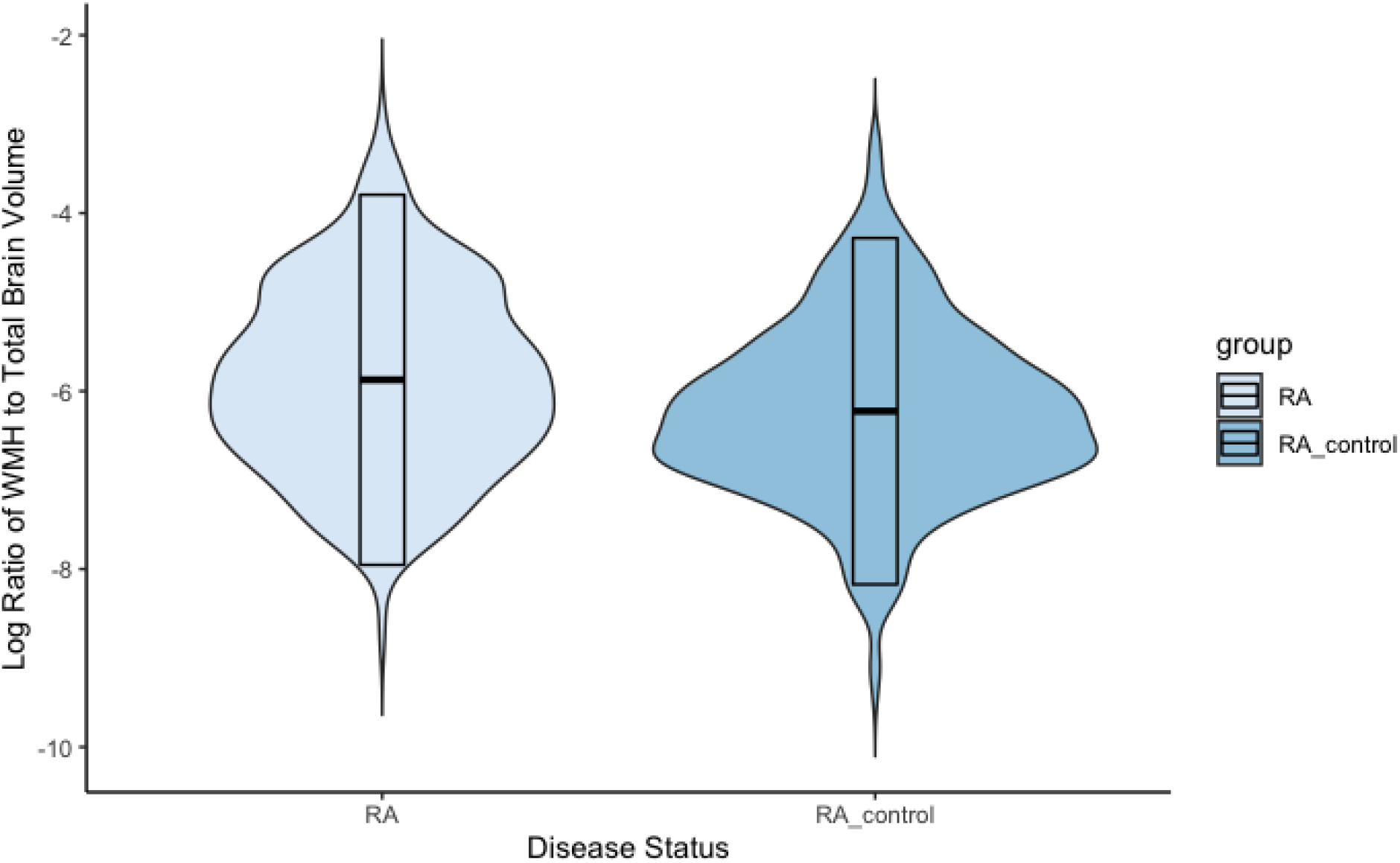
Violin plot representing the log transformed ratio of the total volume of white matter hyperintensities to total brain volume in patients with rheumatoid arthritis as compared to a matched control group.

**Figure 3.**
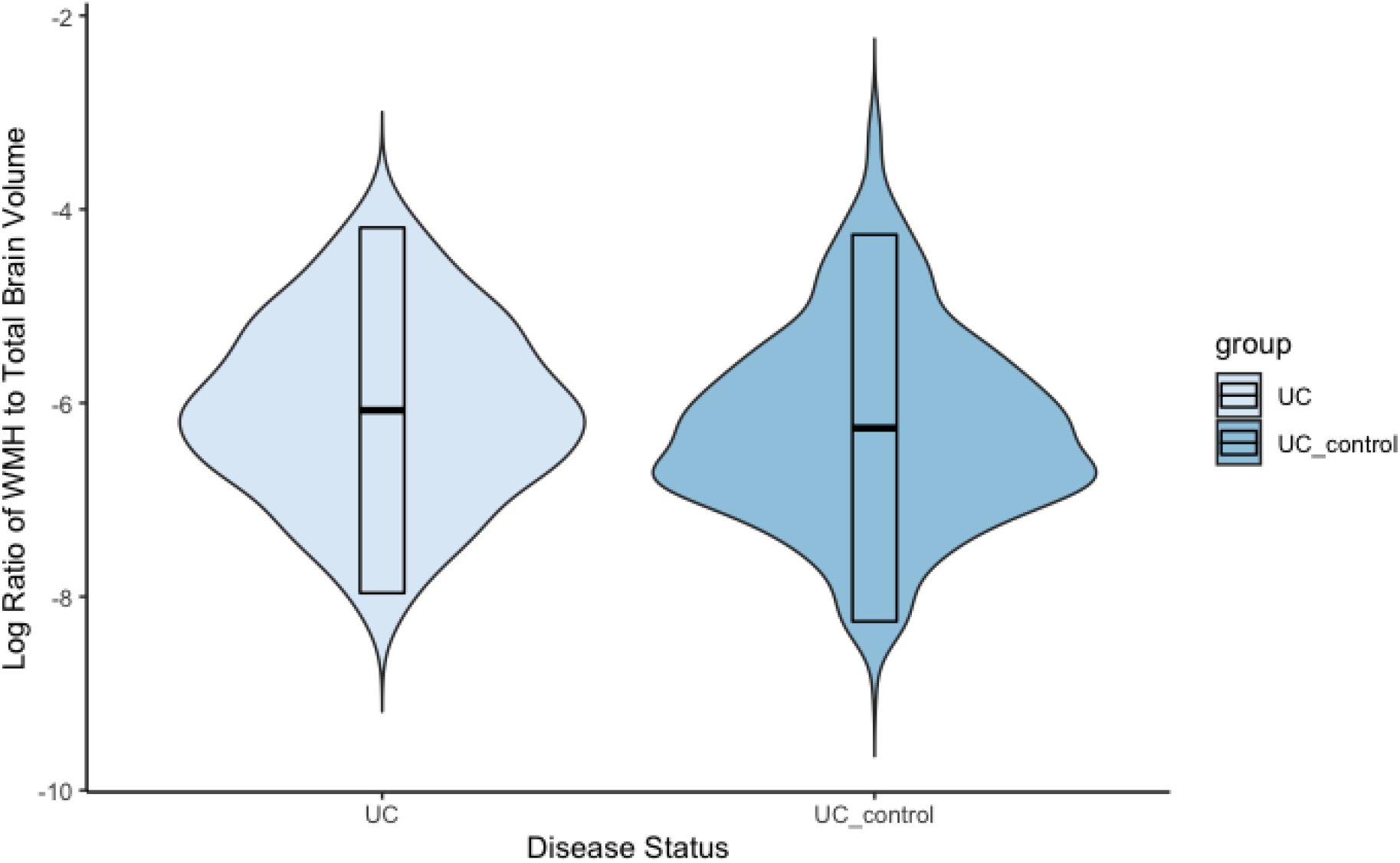
Violin plot representing the log transformed total volume of white matter hyperintensities to total brain volume in patients with ulcerative colitis as compared to a matched control group.

A second model was run including hypertension as a covariate. This was performed to account for any potential signal being attributable to the most prevalent cerebrovascular risk factor in these patient populations. This did account for some of the signal as evidenced by the difference in effect size between models 1 and 2. A similar magnitude of effect difference was seen in both RA and UC between the two models. However, a statistically significant difference was observed in both groups as compared to their controls even when accounting for hypertension. Full results can be found in Table 2 and Table 3.

**Table 2:** Linear model regression results of the log transform of the ratio of total volume of white matter hyperintensities to total brain volume in RA and UC with disease status, age, sex and ICV as covariates

| | Log Transformed<br>Median Ratio of<br>Total WMH to<br>Total Brain<br>Volume Patient<br>Group | Log Transformed<br>Median Ratio of<br>Total WMH to<br>Total Brain<br>Volume Control<br>Group | Model 1<br>p-value | $\beta$ coefficient | 95% CI |
| --- | --- | --- | --- | --- | --- |
| **RA** | **-5.91** | **-6.29** | **$2.0 \times 10^{-8}$** | **-0.36** | **-0.48, -0.23** |
| **UC** | **-6.08** | **-6.33** | **0.003** | **-0.19** | **-0.32, -0.07** |

Significant findings shown in bold.

**Table 3:**
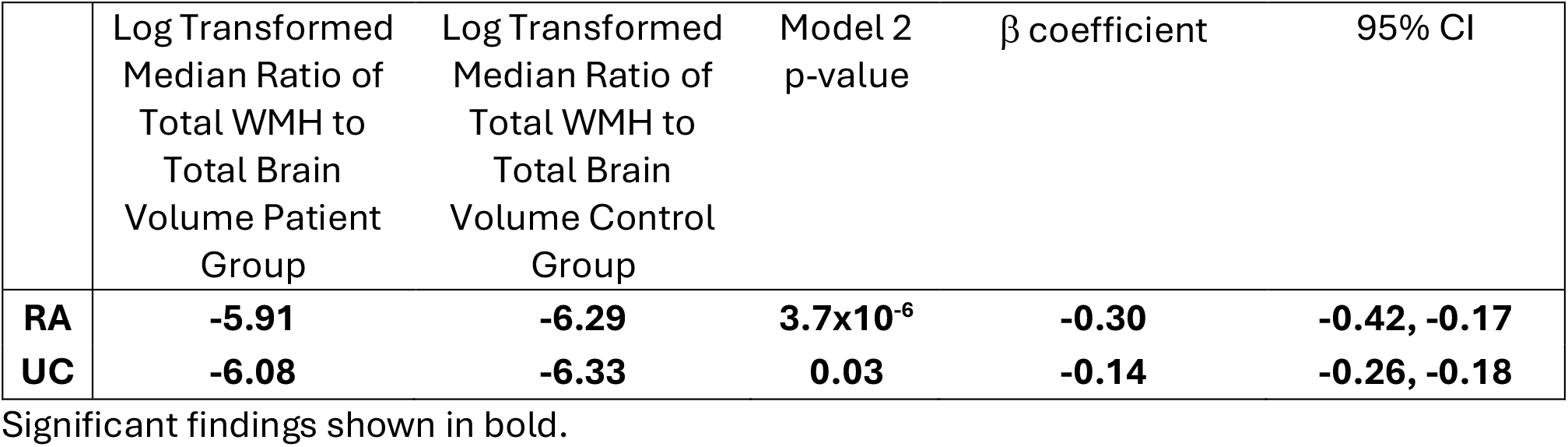
Linear model regression results of the log transform of the ratio of total volume of white matter hyperintensities to total brain volume in RA and UC with disease status, age, sex, ICV, and hypertension as covariates

| | Log Transformed<br>Median Ratio of<br>Total WMH to<br>Total Brain<br>Volume Patient<br>Group | Log Transformed<br>Median Ratio of<br>Total WMH to<br>Total Brain<br>Volume Control<br>Group | Model 2<br>p-value | $\beta$ coefficient | 95% CI |
| --- | --- | --- | --- | --- | --- |
| **RA** | **-5.91** | **-6.29** | **$3.7 \times 10^{-6}$** | **-0.30** | **-0.42, -0.17** |
| **UC** | **-6.08** | **-6.33** | **0.03** | **-0.14** | **-0.26, -0.18** |

Significant findings shown in bold.

### Average lesion distribution mapsa

To qualitatively observe the distribution of these lesions across the brain average lesion maps were created for both the RA and UC patient populations. A similar distribution of lesions was observed in both populations with WMHs being predominantly periventricular. Figure 4 below shows these average maps.

**Figure 4.**
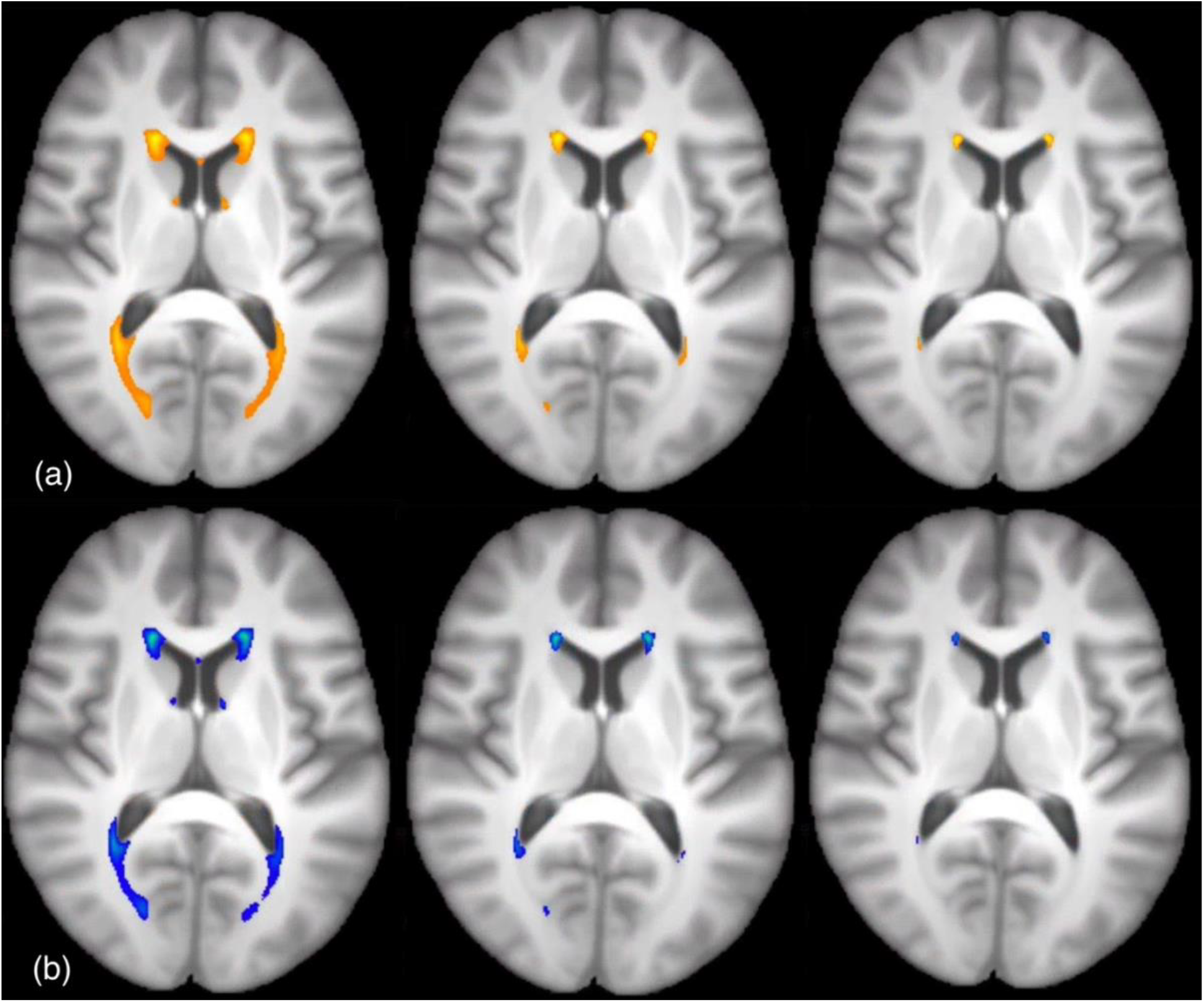
Average lesion maps representing lesion distribution in (a) RA patients (yellow) (b) UC patients (blue) thresholded from left to right at 5%, 50% and 70%.

**Voxelwise statistical analysis of lesion distribution differences**

Voxelwise analysis revealed no significant group differences between RA and UC subjects or between UC subjects compared to their WMH volume matched controls. There was a small but statistically significant cluster seen between RA subjects compared to their WMH volume matched controls.

The Harvard-Oxford atlas returned a non-specific label of “Right Cerebral White Matter”. The JHU atlas returned two regions across the cluster, “Anterior corona radiata R” and “External capsule R”. The result of the threshold free cluster enhancement analysis highlighting this difference is shown below in Figure 5.

**Figure 5:**
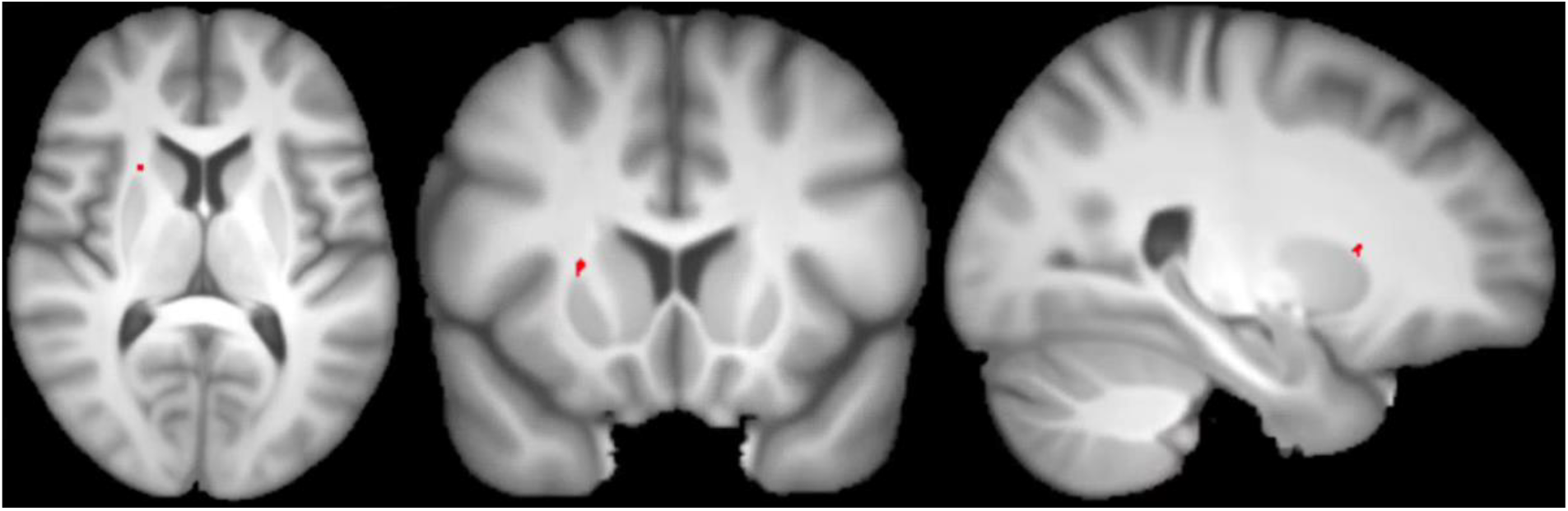
Threshold free cluster enhancement results from voxel wise analysis of RA subjects compared to age, sex and total volume of white matter hyperintensities matched controls. MNI coordinates 24 × 15 × 9.

## Discussion

In the primary analysis, we found a significantly higher volume of WMHs in patients with RA compared to controls. This same result was seen in the UC population to a smaller degree. It’s important to keep in mind that this is a populational data set whereby subjects for inclusion in this analysis were identified using ICD-10 codes. While ICD-10 codes provide a standardized way of classifying diseases it’s well documented that the accuracy of the use of these codes in practice can vary dramatically [32].

Another limitation to this study is the lack of information on disease duration and severity. Both UC and RA are relapsing-remitting diseases and can vary widely in their individual presentation. Previously published data suggests that the age of onset can significantly affect disease severity [33]. How well controlled an individual’s disease is and on what medications may also have an important impact on these central effects which we are unable to quantify with this data set.

As mentioned in the introduction, previous studies that have looked at white matter hyperintensity differences in both RA and UC report varying results. There were important differences in the analytical approach and in the study populations that may have contributed to this difference in results seen between the Bekkelund et al. paper and the Vassilaki et al paper introduced in the beginning [12, 13].Bekkelund et al. used a visual scoring system where the WMHs were identified and the area of the largest lesion was manually estimated as performed by a blinded radiologist, whereas Vassilaki et al used a semi-automated method for segmentation performed by a trained image analyst. In contrast, we used a fully automated segmentation method trained on a curated training dataset. While Bekkelund et al did look at overall total lesion count it did not account for total volume differences as it only looked at the area of the largest identified lesion. The subject population was also significantly smaller with a sample size of 33 patients and 48 controls. The patient population was younger with an average age of 45 and was only comprised of females. Both the Vassilaki et al. study and our current study had a close to 30% male patient and control population. This is in line with epidemiological estimates for gender distribution in RA [34]. This population was slightly younger, however, we reported similar findings to those found by Vassilaki et al. While the Vassilaki et al. study was larger than the Bekkelund et al. study, with a sample size of 104 subjects and used a similar 3:1 matching ratio it was still less than half the sample size of our subject groups which consisted of 250 RA subjects with 756 age and sex matched controls.

Vassilaki et al. also looked at cognitive measures and dementia risk and despite the neuroimaging biomarker differences they did not see a significant difference in incident dementia. This was determined based on an extensive testing regimen of nine neuropsychological tests and then adjudicated by consensus between the study coordinator, physician and neuropsychologist.

Around the same time as the Bekkelund et al. work a report from Geissler et al, came out that looked at lesion presence in both Crohn’s disease and ulcerative colitis subjects compared to controls and reported significantly higher incidence of lesions in both diseases. [35]. This was done from visual reporting alone and was a binary yes or no based on singular presence of WMLs. More recently, Zikou et al. showed a significant difference in the presence of WMHs in IBD patients (including both Crohn’s disease and ulcerative colitis). As with the previous work done this was a small sample of only 18 subjects and was done based on visual counting of the WMHs. This study also found a significant difference in total WMH volume in UC subjects compared to matched controls. This work confirms what was found in these earlier studies but has the added benefit of being a much larger sample size of 240 UC subjects with 720 age and sex matched controls. Additionally, we used more quantitative analysis methods than those utilised in these previous studies.

In the follow-up analysis we matched the control group based on the total white matter hyperintensity volume IDP to look at any differences in the distribution of these lesions. For the UC compared to controls we saw no significant difference in lesion distribution. The same was true when comparing between RA and UC patient groups. Overall, the lack of differences in these various analyses signifies that the increase in WMHs is most likely driven by increased cerebrovascular risk factors. The periventricular distribution of these lesions is in line with previous studies which show an association between the location of WML, periventricular white matter lesions (PVWML) or deep white matter lesions (DWML), and aetiology [36]. PVWML may be more hemodynamically determined and may be more heavily influenced by ischemic and cardiovascular risk factors [37].

There was a small, but significant cluster found in the corona radiata and external capsule in the RA patient population as compared to the control group. This is an area that has been studied extensively in the context of stroke, mild cognitive impairment, dementia, and general cognitive decline [38]. Given the small size of this cluster it is important not to overstate the significance of this finding. However, this does align with previous epidemiological research showing increased risk of cerebrovascular incidents in RA [8, 39-41].

An increased risk of cerebral vascular disease (CVD) and complications such as stroke is well documented in both RA and UC [42-44]. Both this study, and the one conducted by Vassilaki et al., had patients and controls each with similar cardiovascular risk profiles and still found significant differences in these WMH volumes. This suggests that these findings are driven more from a combination of chronic inflammation from these diseases and/or medication toxicity given the known association between disease modifying anti-rheumatic drugs (DMARDS) and WMHs [45, 46].

There is not a large volume of literature looking specifically at WMH volume in these populations. To our knowledge, this is the largest dataset to date to look at WMH volume and distribution in both these diseases. These findings play a potentially important role in further understanding the extent of the effect of inflammation from autoimmune diseases on cerebrovascular health and white matter integrity. This is particularly relevant as we look to better understand the interplay between both disruptions in short-term cognition and cognitive decline and the link between these diseases and future neurodegenerative diseases [47, 48]. This highlights the importance of continued monitoring of cerebrovascular health in people with chronic autoimmune conditions.

## Data Availability

All data produced in the present work are contained in the manuscript.

## Acknowledgements

From the Department of Neuroimaging and the Department of Psychosis Studies, Institute of Psychiatry, Psychology, and Neuroscience, King’s College London

## Statements & Declarations

### Ethics approval and consent to participate

The UK Biobank has approval from the North West Multi-centre Research Ethics Committee (MREC) as a Research Tissue Bank (RTB) approval. This research was done under the RTB approval and separate ethical clearance was not required. The UK Biobank Ethics and Governance Council (EGC) was established as an independent guardian of the UK Biobank Ethics and Governance Framework (EGF). All participant materials, including the informed consent form, have been developed and are monitored under this framework. All data received from the UK Biobank is anonymised and additional consent for this research was not required.

## Data Availability

UK Biobank data are available through a procedure described at https://www.ukbiobank.ac.uk/enable-your-research. All data was accessed under UK Biobank application number 40933.

## Author Contributions

All authors contributed to the study conception and design. Data analysis was performed by Jennifer Cox. The first draft of the manuscript was written by Jennifer Cox and all authors commented on previous versions of the manuscript. All authors read and approved the final manuscript.

